# Molecular markers for early stratification of disease severity and progression in COVID-19

**DOI:** 10.1101/2022.02.06.22270355

**Authors:** Anusha, Savitha Anne Sebastian, N. Raksha, K. Raksha, H. Krishnamurthy, Bhuvana Krishna, George D’Souza, Jyothi Idiculla, Neha Vyas

**Affiliations:** St. John’s Research Institute, Bangalore-560034; St. John’s medical college and hospital, Bangalore-560034; National Centre for Biological Sciences, Bangalore Life Science Cluster, Bangalore-560065; Manipal Academy of Higher Education (MAHE), Manipal 576104,India

## Abstract

COVID-19 infections have imposed immense pressure on the healthcare system of most countries. While the initial studies have identified better therapeutic and diagnostic approaches, the disease severity is still assessed by close monitoring of symptoms by healthcare professionals due to the lack of biomarkers for disease stratification. In this study, we have probed the immune and molecular profiles of COVID-19 patients at 48-hour intervals after hospitalization to identify early markers, if any, of disease progression and severity. Our study reveals that the molecular profiles of patients likely to enter the host-immune response mediated moderate or severe disease progression are distinct even in the early phase of infection when severe symptoms are not yet apparent. Our data from 37 patients suggest that at hospitalization, IL6 (>300pg/ml) and IL8 levels (>200pg/ml) identify cytokine-dependent disease progression. Monitoring their levels will facilitate timely intervention using available immunomodulators or precision medicines in those likely to progress due to cytokine storm and help improve outcomes. Additionally, it will also help identify cytokine-independent progressive patients, not likely to benefit from immuno-modulators or precision drugs.

## Introduction

Coronaviruses are RNA viruses, which belong to the Coronaviridae family and subfamily Coronavirinae. Coronavirinae viruses are known to cause the common cold in humans. Bats or mice act as their natural host, and crossovers to humans are known since 1960(Mittal et al., 2020). In 2002–2003, coronavirus designated as Severe Acute Respiratory Syndrome Coronavirus (SARS-CoV), affected 8422 people in China and Hong Kong and caused 916 deaths (mortality rate of 11%) (Singhal, 2020). In 2012, the Middle East respiratory syndrome coronavirus (MERS-CoV) affected 2494 people and caused 858 deaths (mortality rate of 34%) in Saudi Arabia(Singhal, 2020). In 2019 December, the SARS-CoV2 virus was identified in Wuhan, China, and is responsible for the current pandemic, named COVID-19. SARS-CoV2 is highly contagious and has spread to more than 180 countries. Mutations in the virus have improved its ability to cause the human to human transmission(Wan et al., 2020; Li, 2008). COVID-19 symptoms can range from Lu-like symptoms to severe pneumonia, Acute respiratory distress syndrome (ARDS), clotting disorder as well as multi-organ dysfunction syndrome (MODS), and death. COVID-19 has impacted more than 290 million people worldwide and the numbers are constantly increasing. Healthcare resources and infrastructure have been stretched across the globe.

Clinically, COVID-19 infection triggers a biphasic illness. In the first phase, the virus infects the host and replicates causing, Lu-like symptoms, and most patients recover in 5-6 days without further complications. However, some patients enter the second phase characterised by hyper-inflammation causing pneumonia and or clotting disorders. These complications are due to the virus-induced cytokine storm and cellular and organ damage. The virus-induced cytokine storm followed by hyper-inflammation causes severe pneumonia, ARDS, or MODS which can lead to death if not treated appropriately and timely. India is currently grappling with the third wave of COVID-19, and as of 4 Jan 2022, there are ∼35 million confirmed cases with around 1.3% deaths. The current therapeutic approach can cure many severe patients even with >50% lung damage, if medical intervention is timely. While reliable tests are available for rapid and early detection of SARS-CoV-2 infection; there are no reliable tests available that can anticipate the second phase of the illness. Currently, this is addressed by close monitoring for onset of the symptoms like breathlessness or deterioration in oxygen saturation by which time it may already be too late to intervene. This study was designed to address this urgent need. Here we attempted to identify early biomarkers for patients likely to enter the more serious second phase of the disease.

In the first part of the study, we evaluated the temporal profile of 9 serum markers (cytokines and lung markers), lymphocytes, and neutrophils in 14-patients on the day of hospitalization (denoted as Day0 i.e. day3-7 after the onset of symptoms) and at 48hrs interval during their hospital stay, while the clinical course was also monitored. We found that the number of B cells and T cells subtypes show variability with time but do not show a significant difference between mild non-progressive and progressive patients. On the other hand, most cytokines and lung markers tested showed differential levels in mild versus severe disease. Based on this preliminary data, we selected 5 serum markers for analysis in a larger number of patients (37 patients), on Day0.

Our study reveals that serum levels of IL6, IL8, and Surfactant protein-D (SP-D) can be used as early markers to stratify the severity of COVID-19, even on the day of hospitalization and before hypoxia sets-in. IL6 and IL8 together can identify ∼50% of patients with moderate or severe COVID-19 progression even when the severe symptoms of the disease progression are not yet apparent. This can aid in well-timed therapeutic intervention with available immuno-modulators or precision medicine. Importantly, these markers can also identify patients who are progressive even in absence of hyper-inflammatory response and are not likely to benefit from the immuno-modulators or precision drugs like Tocilizumab.

## Results

### NLR-ratio, T-cells, or B-cells are not different in progressive COVID-19 patients

We recruited a total of 42 patients in this study, 5 patients with alcoholic liver disease or HIV were excluded from molecular analysis as these conditions are known to influence the molecular profiles of interest independently from SARS-CoV2 infection or severity status (Hong et al., 2002; Kawaratani et al., 2013)(Figure 1A, Supplementary Table 1). In the pilot stage of the study, venous blood and serum were collected from 14-patients for a time-course analysis of the molecular players of interest. This allowed us to map the temporal profile of the markers of choice and identify representative early molecular markers of progressive COVID-19. In the second stage of the study, only serum samples were collected on the day of admission (Day 0) to identify trends in a larger number of patients for necessary statistical significance (Figure 1A). Molecular data were analysed and correlated retrospectively with the clinical data. Clinically, patients are divided into three groups based on the percent saturation of the oxygen (SpO2): mild (SpO2 >94%), moderate (SpO2 93-90%), or severe (SpO2 <90%) (Supplementary Figure 1). The lowest SpO2 levels during the stay is considered for the clinical classification.

**Figure 1:**
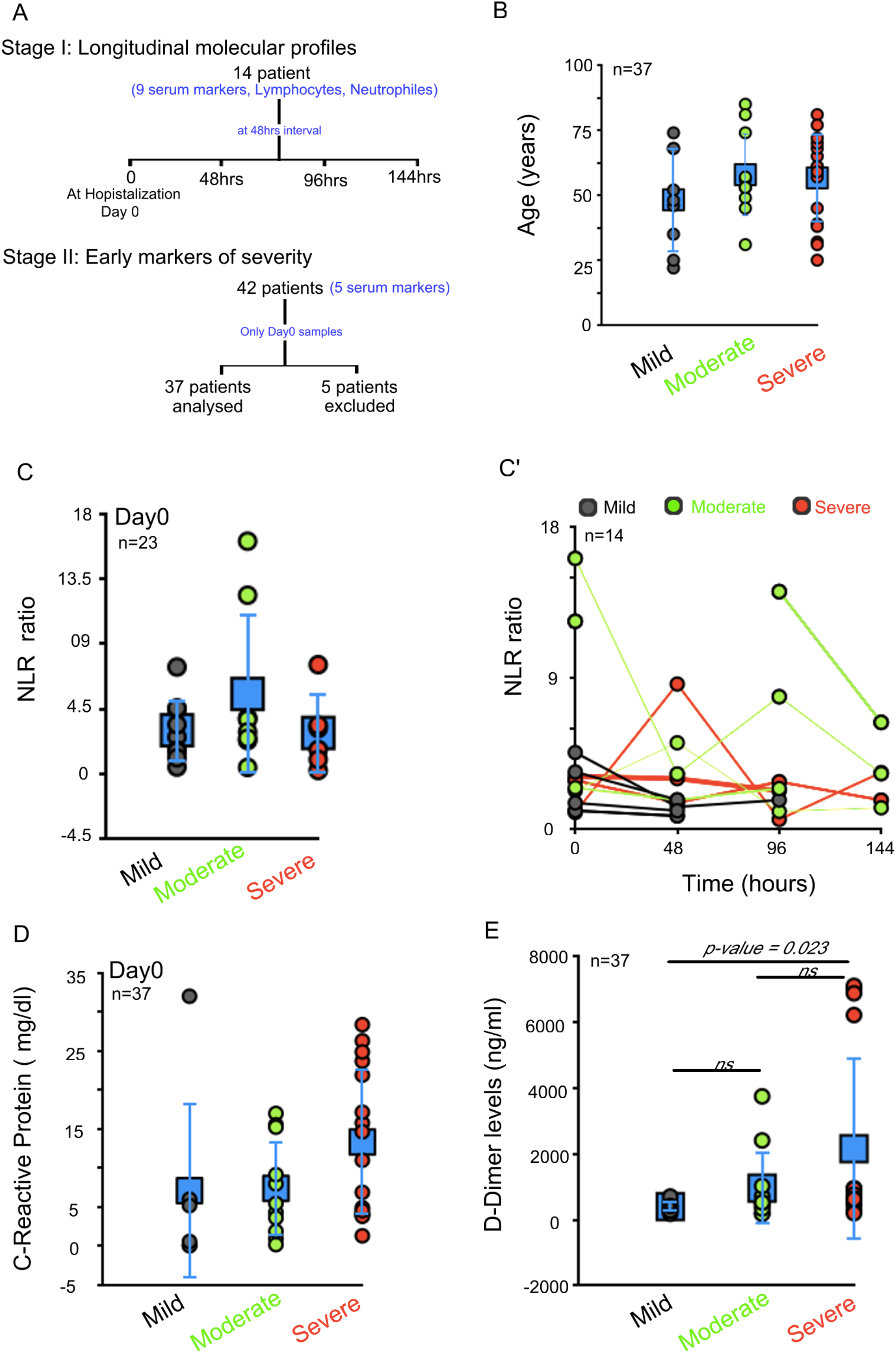
NLR and CRP fails to predict disease severity in COVID-19 patients. A) Flow chart representing the study design. B) Dot plot represents age of the patients included in the study. C) Dot plot represents distribution of neutrophils to leukocyte ratio (NLR) in patients at the time of admission. C’) Line graph represents time course analysis of NLR, at 48 hr interval, FACS based analysis D) Dot plot represents levels of C-reactive proteins (CRP, mg/dl) among COVID-19 patients with varying severity on Day0. D) Dot plot represents levels of d-Dimer among COVID-19 patients with varying severity. Grey, mild COVID-19; Green, moderate COVID-19; Red, severe COVID-19 patients. Day 0 is the hospital admission day. Blue data point, average ± S.D. across patients in the group.

We first analyzed the known markers for predicting the severity of the disease, which includes neutrophil to lymphocyte ratio(NLR;)(Zhang et al., 2020; Ponti et al., 2020; Zhu et al., 2020), d-Dimer(Ponti et al., 2020; Zhu et al., 2020; He et al., 2021) and c-Reactive protein (CRP) levels (Ponti et al., 2020). The patients in our study were in the range of 20-80years, disease severity was independent of the age (Figure 1B, Supplementary Table 1). NLR was not different between mild, moderate, or severe disease at the time of admission (Figure 1C) or during their hospitalization phase (Figure 1C’). C-reactive protein (CRP) levels, a popular inflammatory marker, were also similar between mild (mean value 7.06±11.32mg/dl), moderate (7.36±6.12mg/dl), and severe disease (13.30±9.43mg/dl) for its reliable clinical use (Figure 1D). d-Dimer levels were different between the patients with mild (395.429±179.24ng/ml), the moderate (954.81±1111.24ng/ml), and the severe (2150±2778ng/ml) disease. While statistical significance can be identified between the mild vs the severe disease for d-Dimer levels (p-value = 0.023, Figure 1E); the spread in the values of d-Dimer can make interpretation of individual cases difficult for necessary decisive action at clinics.

Earlier studies have highlighted that lymphopenia is a consistent phenotype in most Covid-19 patients(Cao, 2020; Tavakolpour et al., 2020). Lymphocytes levels (mostly T-cells) were significantly lower in patients with severe vs mild disease and are thought to be useful as an early marker of disease severity(Zhang et al., 2020; Qin et al., 2020). We hence reviewed the levels of T and B-lymphocytes in our study. We assessed the T cell subsets of CD8+ T-cells (CD45+ CD3+ CD8+), and helper T-cells (CD45+ CD3+ CD4+), by evaluating a standard combination of cell surface markers via FACS based analysis. There was no significant difference in CD4+ or CD8+ T-cells between the various severity groups on the day of hospital admission (Figure 2A-B). Even in time-course analysis, at 48hrs across their hospitalization period, there is no significant difference in the T-cell subsets between patients with severe, moderate or mild disease progression (Figure 2A’-B’). We analyzed the cell surface markers of the B-cell population via FACS based analysis. B-cells were divided into, regulatory B-cells (CD45+ CD19+ CD24+ CD38+), memory B-cells (CD45+ CD19+ CD24+ CD38-), and naïve B-cells (CD45+ CD19+ CD24-CD38-). B-cell subtypes also did not show any significant difference between the various severity groups on day0 (Figure 3A-C) or in the time-course analysis (Figure 3A’-C’). We hence conclude that T-cells or B-cells are not substantially different in patients with mild or progressive COVID-19 disease.

**Figure 2:**
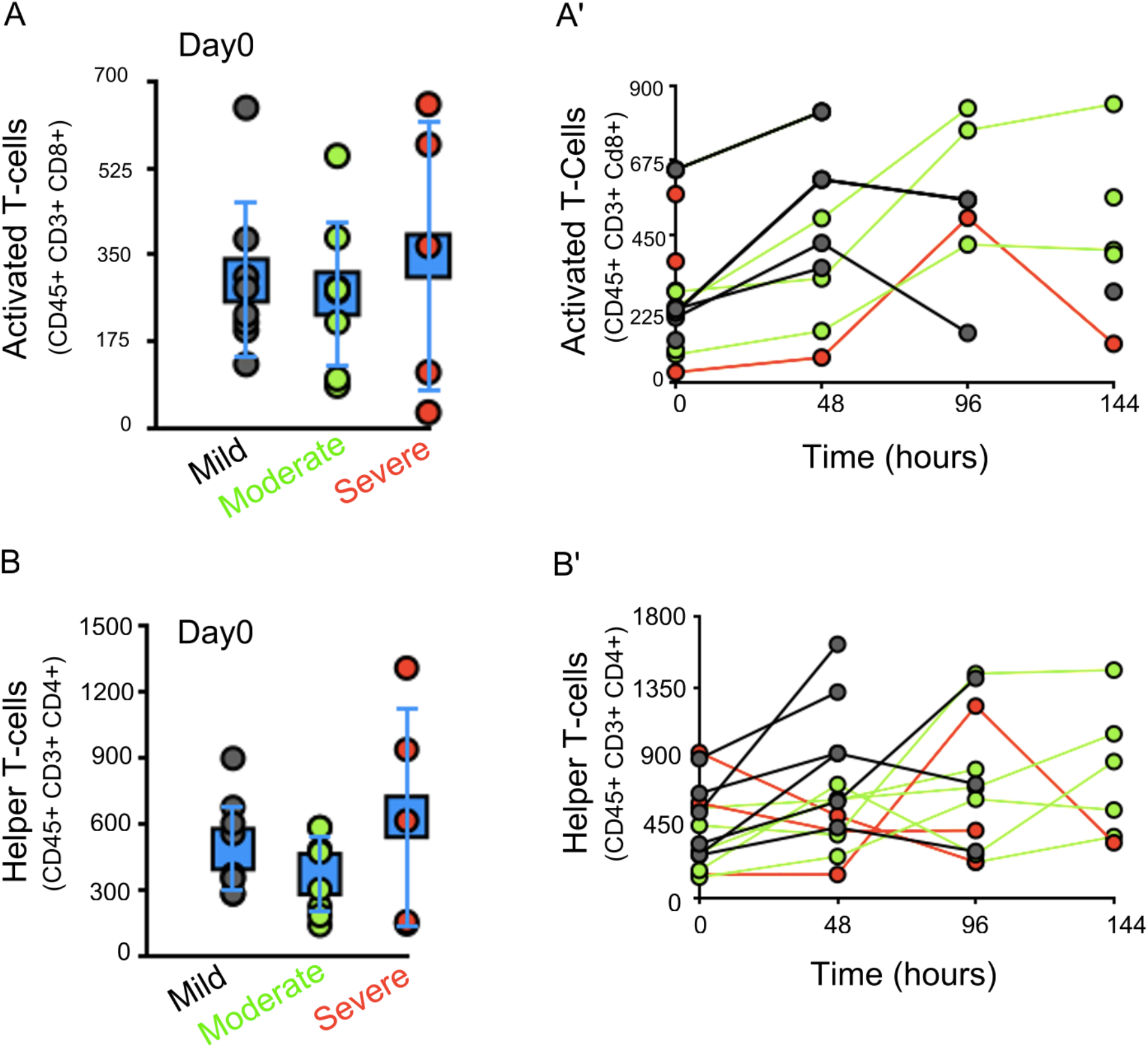
T-cell numbers are not different in COVID-19 patients with varying disease severity. A) Dot plot represents FACS based analysis of Activated T-cells (CD45+ CD3+ CD8+) in peripheral blood, on Day0, in patients with varying severity. A’) Time course analysis of Activated T-cell profile in peripheral blood in patients with varying severity, at 48hr interval. B) Dot plot represents FACS based analysis of Helper T-cells (CD45+ CD3+ CD4+) in peripheral blood, on Day0, in patients with varying severity. B’) Time course analysis of Helper T-cells in peripheral blood in patients with varying severity, at 48hr interval. Blue data point in A, B and C represents, average ± S.D. Grey, mild COVID-19; Green, moderate COVID-19; Red, severe COVID-19 patients, Day 0 is the hospital admission day.

**Figure 3:**
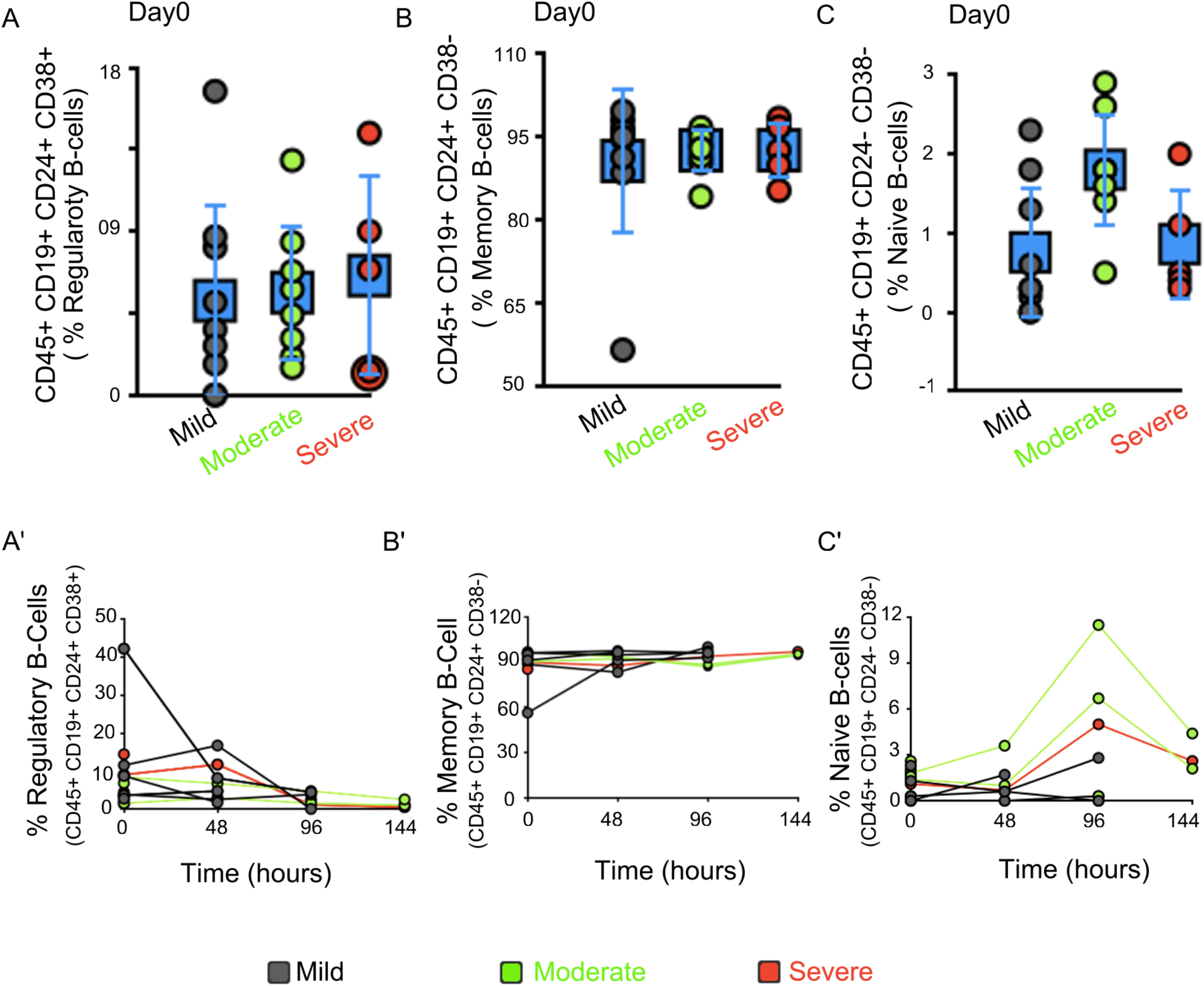
B-cell numbers are not different in progressive COVID-19 patients. A) Dot plot represents percentage of regulatory B-cells (CD45+ CD19+ CD24+ CD38+) in peripheral blood, on Day0, in patients with varying severity. A’) Line graph represent changes in percentage of Regulatory B-cells in peripheral blood of patients in patients with varying severity, at 48hr interval. B) Dot plot represents FACS based analysis of percentage of memory B-cells (CD45+ CD19+ CD24+ CD38-) in peripheral blood, on Day0, in patients with varying severity. B’) Line graph represent changes in percentage of memory B-cells in patients with varying severity, at 48hr interval. C) Dot plot represents percentage of naive B-cells (CD45+ CD19+ CD24-CD38-) on Day0, in patients with varying severity. C’) Line graph represent changes in percentage of naive B-cells in patients with varying severity, at 48hr interval. Blue data point in A, B and C represents, average ± S.D. Grey, mild COVID-19; Green, moderate COVID-19; Red, severe COVID-19 patients. Day0 is the hospital admission day.

### Severe patients show upregulation in most Interleukins levels tested

We evaluated the cytokines in the serum of these patients to understand their temporal profiles. IL-8 levels were remarkably different even on Day0 and during the hospitalization phase in patients with severe (626.53±29.70 pg/ml) versus mild disease (203.48±113.53 pg/ml) (Figure 4A), unlike moderate disease (331.47±210.34 pg/ml; Figure 4A).

**Figure 4:**
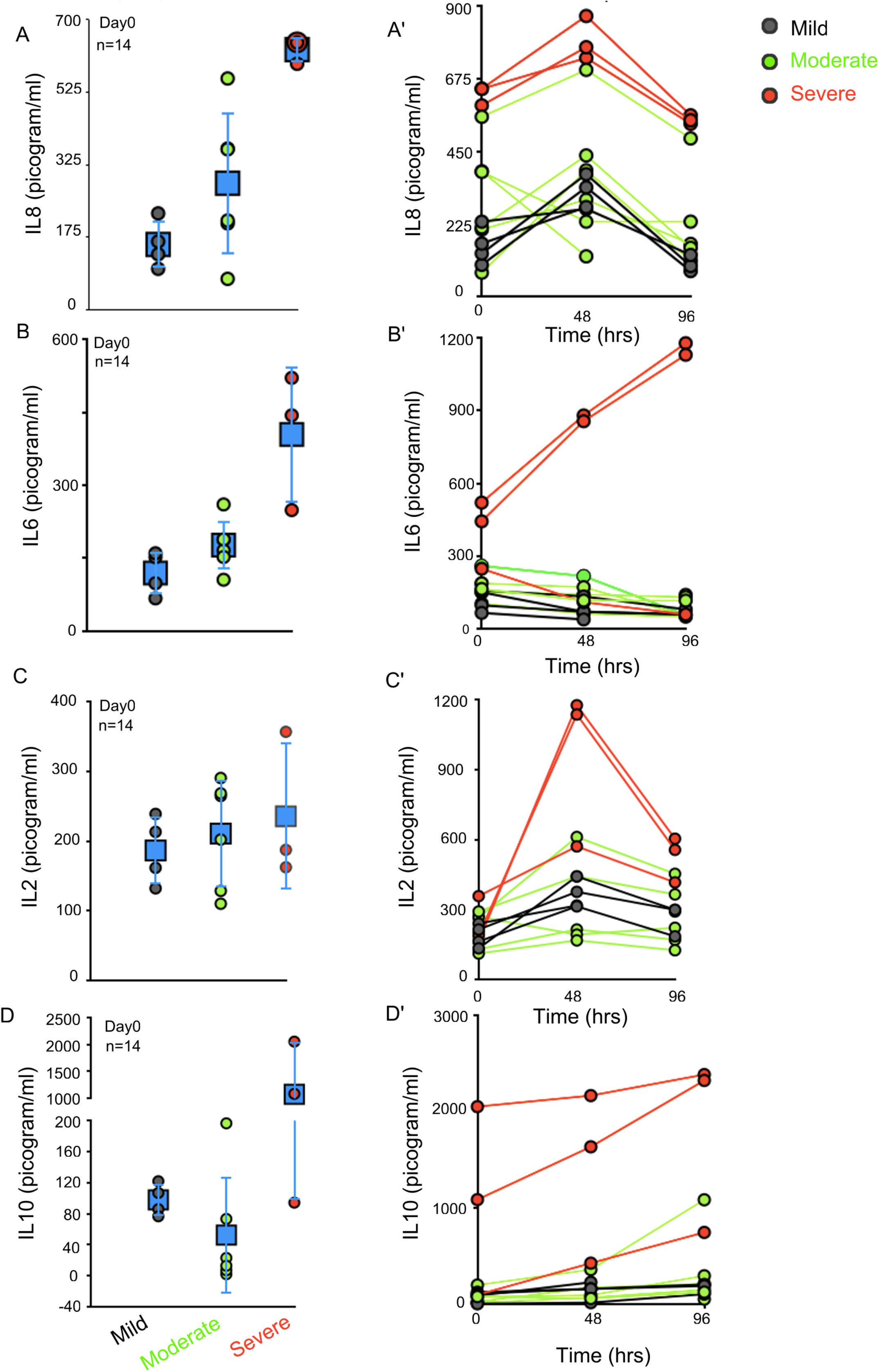
IL2, IL6, IL10 and IL8 are elevated in severe COVID-19 patients. A-D) Dot plot represents ELISA based analysis of serum levels of IL8 (A), IL6 (B), IL2 (C) and IL10 (D), on Day0, in patients with varying severity. A’-D’) Line graph represents changes in serum IL8 (A’), IL6 (B’), IL2 (C’) and IL10 (D’), levels in patients, at 48hr interval. IL6 levels are significantly upregulated in some patients even on Day0. IL8 levels show large variability in moderate patients but are elevated in some severe patients on Day0. IL2 levels show large spread in values across disease severity. IL10 levels are upregulated in some severe patients unlike mild or moderate patients. Blue data point in A, B and C represents, average ± S.D. Grey, mild COVID-19; Green, moderate COVID-19; Red, severe COVID-19 patients. Day0 is the hospital admission day.

IL6 levels were also high in 2 out of 3 patients with severe disease tested in our pilot study (404.75±140.31 pg/ml) compared to patients with mild disease (119.40±44.32 pg/ml). The IL6 levels show further elevation during the hospitalization phase in 2 patients with severe disease (1153.83±33.91pg/ml, at 96hrs). While the patients with severe disease but low IL6 level at hospitalization, shows further decrease in IL6 levels with time (Compare red line graphs, Figure 4B’), suggesting an IL6-independent disease progression.

At the time of admission, IL2 levels were not significantly different between the various groups of patients (Figure 4C; mild=186.32±48.38 pg/ml, moderate=210.54±76.84pg/ml or severe= 235.31±105.67 pg/ml). IL2 increases significantly, but only transiently in 2 of the 3 patients with severe disease (Figure 4C’, 1156.35±28.09pg/ml) at 48hrs post-admission compared to patients with mild disease (Figure 4C’, 361.088±60.55pg/ml). IL10 levels were also significantly elevated in 2 out of 3 patients with severe disease (1564±689.35pg/ml) versus mild disease (97.74±20.41pg/ml; compare Figure 4D and 4D’); while IL4 levels remain similar on Day0 and in the time-course analysis for all 14-patients tested (Supplementary Figure 2; 200-350pg/ml). TNF-α levels were below detection level in all the patients tested.

### Lung biomarker, SP-D, can act as an early marker of disease severity

As COVID-19 is known to involve lung damage, we decided to evaluate the temporal profile of lung biomarkers in these patients. We used three lung markers for this study, which include Clara-Cell protein-16 (CC-16), also known as Uteroglobin; Surfactant Protein-D (SP-D); and Secretory Receptor for Advanced Glycation End product (sRAGE). CC-16 is higher in patients with severe disease (48.41±2.77 ng/ml, red data-point, Figure 5A) compared to the mild disease (21.89±13.08 ng/ml, grey data-points, Figure 5A) but only on Day0 (compare Figure 5A and 5A’). Unlike CC-16, SP-D levels are significantly elevated in 2 out of 3 patients with severe disease on Day0 (139.79±12.59 ng/ml) versus mild (19±13.3 ng/ml) or moderate disease (15.72±7.83 ng/ml) (Figure 5B). SP-D levels remain high in 2 out of 3 patients with severe disease during the hospitalization period (Figure 5B’). sRAGE on the other hand shows a diametric profile. Serum levels of sRAGE are higher in mild disease with normal lungs (Figure 5C, 7789.23±2462.78 pg/ml) on the day of hospitalization compared to severe disease (3255.11±1371.09 pg/ml) (Figure 5C). sRAGE levels decrease with time (Figure 5C’), suggesting the differential ability of the infection-induced upregulation of sRAGE in most patients, also known for its anti-inflammatory role(Nakashima et al., 2010).

**Figure 5:**
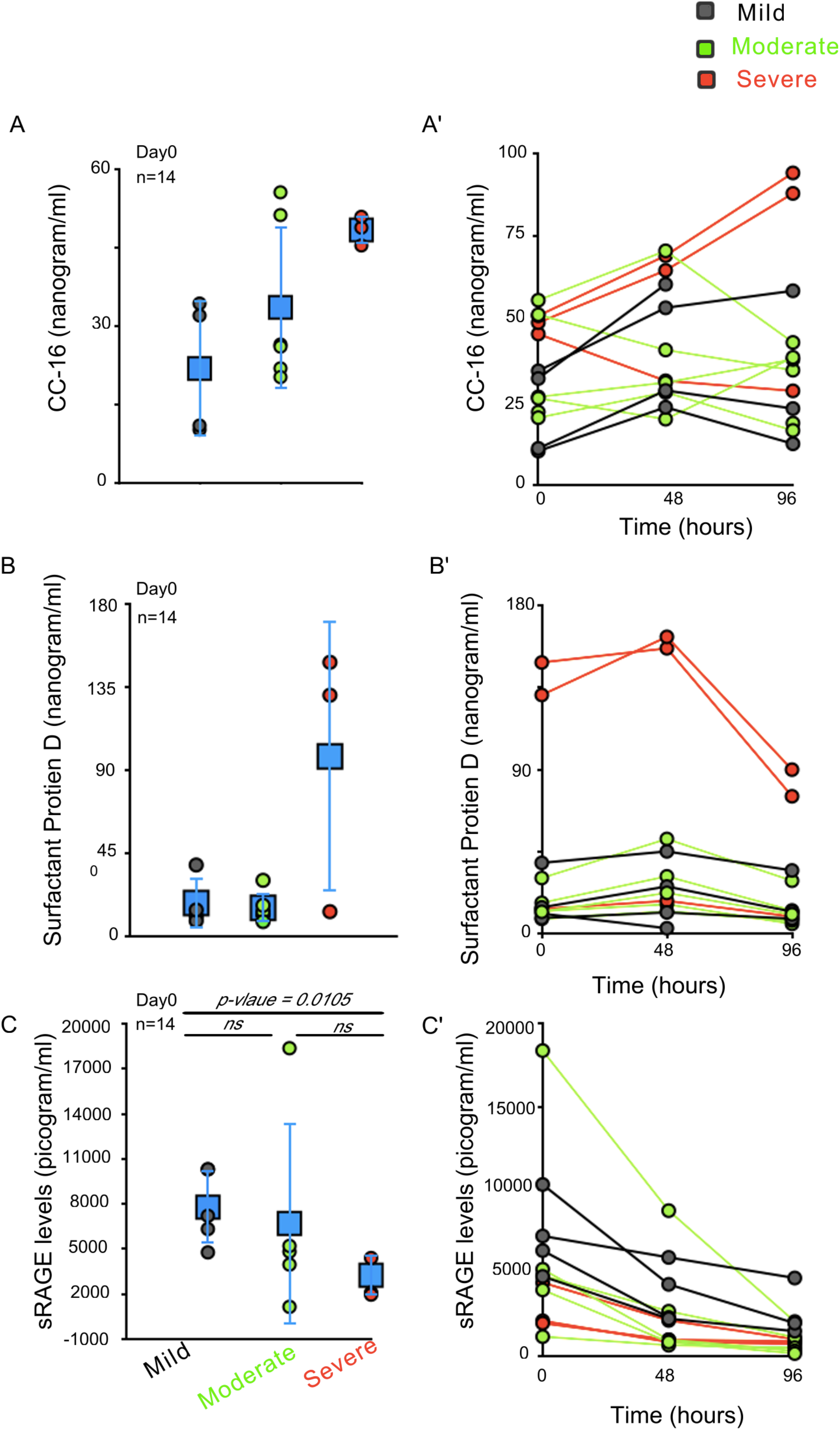
SP-D is upregulated in severe COVID-19 patients unlike sRAGE. A-C) Dot plot represents ELISA based analysis of serum levels of CC-16 (A); SP-D (B) and sRAGE (C), on Day0, in patients with varying severity. A’-C’) Line graph represents changes in serum CC-16 (A’), SP-D (B’) and sRAGE (C’) levels in patients, at 48hr interval. Serum CC-16 levels are elevated in some moderate and all severe patients, on Day0. SP-D levels are elevated in some severe patients on Day0. sRAGE levels are elevated in mild patients compared to severe patients on Day0 only. Blue data point in A, B and C represents, average ± S.D. Grey, mild COVID-19; Green, moderate COVID-19; Red, severe COVID-19 patients. Day 0 is the hospital admission day.

Based on the finding from our temporal pilot study, we identified 5 serum markers namely IL6, IL8, SP-D, CC-16, and sRAGE, for analysis in a larger number of patients to evaluate their reliability as early markers of progressive disease. These markers were evaluated in a total of 37 patients, now only on Day0. We find that both IL6 and IL8 were significantly upregulated in patients with severe disease compared to mild disease (Figure 6A and 6B). IL6 levels are not different between mild (130.47±38.22 pg/ml), and moderate disease (159.38±48.73 pg/ml), but vary significantly in mild (130.47±38.22 pg/ml) versus severe disease (349.69±181.98, p-value= 0.00022, Figure 6A) and between moderate and severe disease (Figure 6A, p-value = 0.00087).

**Figure 6:**
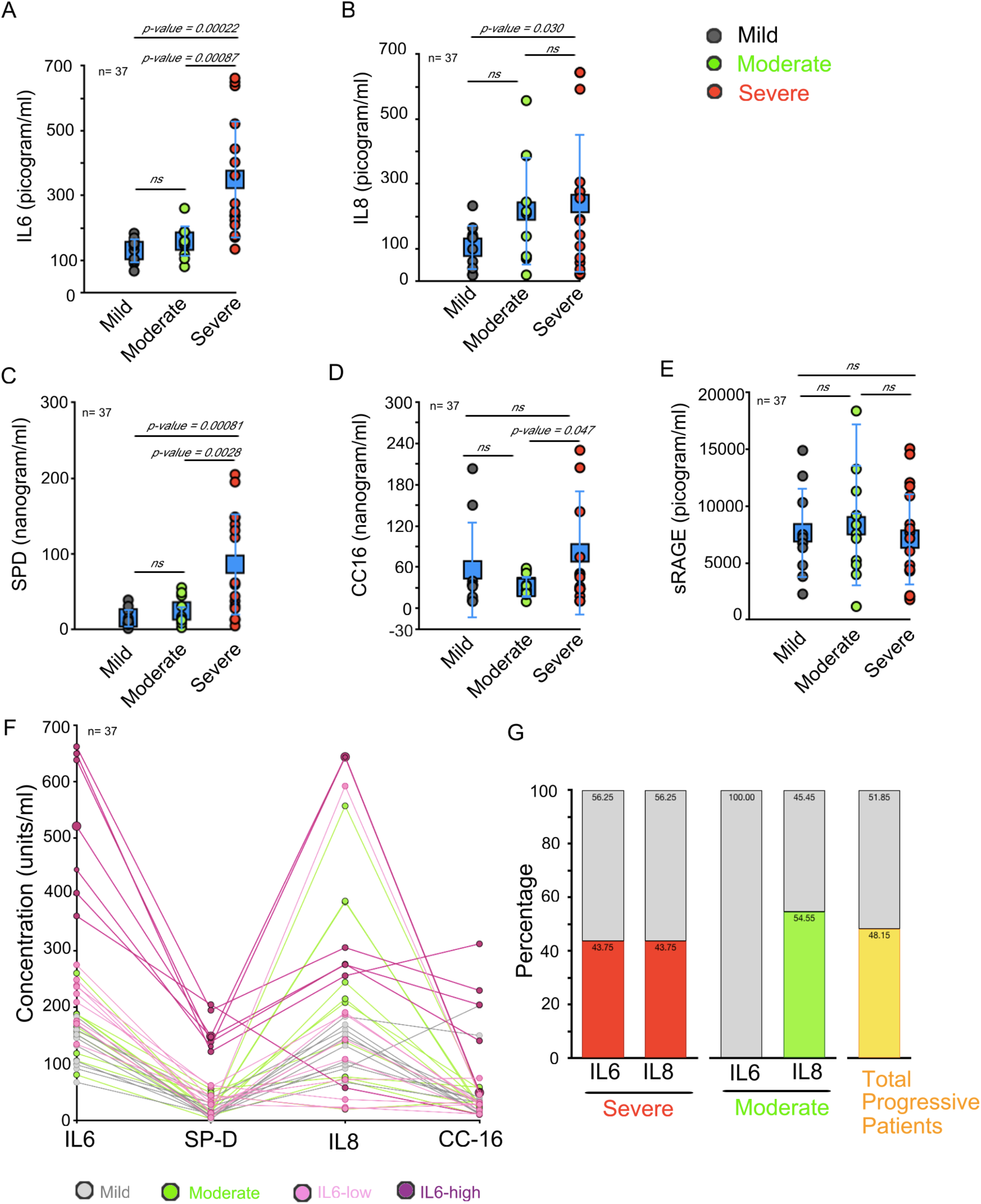
IL6 and IL8 can be used as early markers of progressive COVID-19 disease. A-E) Dot plot represents ELISA based analysis of serum levels of IL6 (A); IL8 (B) SP-D (C), CC-16 (D) and sRAGE (E) on Day0, in COVID-19 patients. F) Line graph represents changes in serum levels of IL6, IL8, SP-D and CC-16 per patient. Light-pink lines represent patients with IL6-low (300pg/ml), dark-pink lines represent patients with IL6-high (>300pg/ml), grey lines represents mild and green-lines represents moderate COVID-19 patients. G) Bar-graph representing percentage of COVID-19 patients identified with IL6-high (>300pg/ml) or IL8-high (>200pg/ml) in severe patients (red bars) or Moderate (green bars) category, yellow bar represents percentage of total patients that fall in the progressive category with IL-high(>300pg/ml) or IL8-high (>200pg/ml) levels. Grey bars represents patients in each category with Il6-low (<300pg/ml) or IL8-low (<200pg/ml) levels. Blue data points (square) in A-E represents, average ± S.D. Grey, mild COVID-19; Green, moderate COVID-19; Red, severe COVID-19 patients. Day0 is the hospital admission day.

IL8 levels show significant difference only between mild (104.92±69.61 pg/ml) and severe disease (239.57±214.05 pg/ml; Figure 6B, p-value = 0.030). We observed that patients with severe (239.57±214.05 pg/ml, Figure 6B) and moderate disease (216.35±167.01 pg/ml, Figure 6B) show a higher spread in serum IL8-levels. Higher variability between IL6 and IL8 might suggest a necessary molecular stratification of the progressive patients into “cytokine-dependent”, and “cytokine-independent” categories.

Further of the three lung markers evaluated, serum SP-D levels are significantly up-regulated in patients with severe (86.25±67.74 ng/ml) compared to mild disease (15.34±12.19 ng/ml, Figure 6C, p-value = 0.00081). SP-D levels for moderate disease (24.57±18.86ng/ml) are not significantly different from mild; unlike severe disease (Figure 6C, p-value= 0.0028).

Again, CC-16 serum levels show significant spread, rather than differences between mild (56.16±70.28ng/ml), moderate (30.44±15.59 ng/ml), and severe disease (80.79±91.36 ng/ml) (Figure 6D). While sRAGE failed to show any significant difference between mild, moderate, or severe disease conditions (Figure 6E). Given the large dynamic range of the markers tested between the clinically defined mild, moderate and severe categories of patients, we decided to evaluate their relative levels and trends per patient (Figure 6F). For this, we divided the severe patients into IL6-low (<300 pg/ml, light pink line-graphs, Figure 6F) and IL6-high (>300 pg/ml, dark pink line-graphs, Figure 6F). This analysis reveals that all IL-6-high patients show upregulation in SP-D, IL8, and CC-16 levels. The IL6-low (<300 pg/ml) but clinically severe patients have IL8, SP-D, or CC-16 levels similar to the mild disease. Interestingly, clinically moderate disease show upregulation in IL8-levels (>200pg/ml) but not in SP-D, CC-16, or IL6 levels (green line-graphs, Figure 6F). We hence propose that IL6 upregulation is associated with higher inflammatory status and damage in COVID-19 patients. Only IL8-high (>200 pg/ml) levels might serve as marker of cytokine-induced moderate disease. Our data also indicate that, IL8-upregulation alone can lead to progressive disease, but the extent of inflammation-induced damage is less in these patients. Using >300 pg/ml as the cut-off for IL6 and >200pg/ml IL8 in our study, 50% of the patients with progressive (moderate or severe) COVID-19 can be identified for early therapeutic intervention (Figure 6G). Importantly our data also pin-points that 50% of the patients in our study had rather cytokine-independent progression.

We propose that only patients with IL6-high (>300pg/ml) should be considered for early intervention with the available IL6-inhibitor drugs like Tocilizumab. Also, not only Tocilizumab but even immuno-modulators should be avoided in patients with IL6-low (<300pg/ml) and IL8-low levels (<200pg/ml) as they might be progressive due to the direct influence of the virus, independent of host-immune responses.

## Discussion

COVID-19 infection has challenged the health care system globally for more than two years now. The initial viral genome sequencing efforts have identified that there are several mutations in the receptor-binding domain (RBD) of the Spike (S)-proteins of the SARS-CoV2. These mutations make this COVID-19 causing virus highly infective by increasing affinity for the ACE-2 receptors(Yi et al., 2020). SARS-CoV and the SARS-CoV2 viruses use the same S-protein to infect the host cells(Yi et al., 2020; Gui et al., 2017; Wu et al., 2012) yet, they do not show significant antibody cross-reactivity. This has been attributed to the mutations in the antigenic epitopes of the S-protein with 92.7% unique epitopes contributed by the non-conserved region. SARS-CoV-2 has 20 unique epitopes as compared to SARS-CoV in the S-protein region and 40 epitopes from SARS-CoV are not present in SARS-COV-2, only 5 epitopes are shared(Zheng and Song, 2020; Mittal et al., 2020). Due to this, SARS-CoV-specific antibodies do not bind to the S-protein of SARS-CoV-2 and there is a need to develop SARS-CoV-2 related antibodies and vaccines independently. Currently, it is expected that available COVID-19 vaccines may not prevent the infections altogether but can mitigate or block lung involvement in most patients(Hodgson et al., 2021). Vaccine manufacturing to cater to the entire population has been challenging for most countries and is turning out to be the current rate-limiting step in allowing us to resume the requisite socioeconomic normalcy. One of the challenges across the globe has been the flooding of hospitals with COVID-19 patients and shortage of critical care beds for the sick. Hence identifying additional avenues to intervene post-infection will be needed to tackle this pandemic. Knowledge of who is likely to progress to severe disease will help triage patients better. Similarly identifying patients early in the course of disease who will benefit from immunomodulators or newer therapies will help early intervention and better outcomes. So far this has been done using clinical characteristics which may be too late as in degree of hypoxia or not very discriminatory like age, presence of comorbidities.

Many COVID-19 related studies done in early 2020 have improved the understanding of the SARS-CoV-2 virus and have facilitated the rapid development of therapeutic interventions. However, several molecular aspects about COVID-19 infection and disease severity remain poorly resolved. Severe COVID-19 is characterized by hyper-inflammation leading to ‘cytokine storm’ which results in widespread inflammatory damage particularly to the lung. We hence studied select cytokines and lung injury markers to evaluate if they can anticipate COVID-19 patients progressing to severe disease. We evaluated serum levels of select cytokines, and lung markers at 48 hourly intervals from the time of admission through the duration of hospitalization at our tertiary care center. Our study has identified early biomarkers that can pinpoint COVID-19 patients likely to progress to severe disease triggered by host-immune responses. Such patients could be considered for timely therapeutic intervention with available immuno-modulator drugs or precision medicines.

Interleukins (ILs) can act as autocrine and paracrine pro-inflammatory or anti-inflammatory molecules. ILs and most other cytokine levels in our body are highly dynamic and increase in several pathological conditions(Dinarello, 2007). Several cells are involved in cytokine secretions, these include, fibroblasts, keratinocytes, vascular endothelial cells, pericytes, mast cells, macrophages, dendritic cells, and T and B cells. Cytokine-storm is a term coined to describe the secretion of high levels of several cytokines that can increase inflammation(Fajgenbaum and June, 2020). SARS-CoV2 virus-induced host immune responses can induce lung damage, thrombosis, multi-organ failure, and death in COVID-19 patients(Fajgenbaum and June, 2020; Mustafa et al., 2020; Hojyo et al., 2020). Currently, clinics are using immuno-modulators or precision drugs only if and when severe symptoms appear. Our data suggest that progressive patients can be identified even in the early phase of the disease for timely intervention with available immuno-modulators. In our study, ∼50% of severe patients show upregulation of most of the ILs tested (accept IL4), much before the manifestations of severe clinical phenotypes. We find that patients with high IL6-levels (>300pg/ml) at hospitalization, are distinct and may be wired for a severe manifestation of the disease. Progressive COVID-19 condition caused mainly by cytokine-storm, if identified early, and treated in-time, can prevent damage to vital organs and improve outcomes. There are several inhibitor drugs available to treat cytokine storm, including ACE inhibitors and Angiotensin II Receptor Blockers, and Corticosteroids. The inhibitor of IL6, Tocilizumab, has also been explored as a therapeutic agent to overcome cytokine-storm in severe COVID-19 patients with varying outcomes(Salama et al., 2020; Salvarani et al., 2021).We suggest that severe patients should be further stratified based on their IL6 and IL8 levels, into the “cytokine-dependent or “cytokine-independent” categories. Immuno-modulators or precision drugs must be restricted to the “cytokine-dependent” category of patients thus obviating potential side effects of these drugs.

However, ∼50% of patients show cytokine-independent progression (Figure 6F and 6G). Severe patients with low levels of IL6 (<300pg/ml) do not show upregulation in IL8, CC-16, or SP-D levels in our study. We hence propose that these patients are progressive without the inflammatory host-immune response or cytokine-storm and are not be suitable for immuno-modulators or precision medicine-based intervention. Also, moderate patients often do not show upregulation in IL6 levels but can have upregulated IL8 levels only (Figure 6F and 6G). Such patients should not be given IL6-inhibitor drugs like Tocilizumab.

The selective upregulation of IL8 in moderate patients suggests that secretion of IL6 and IL8 are independently regulated. Molecules like Trek-1 (a potassium channel) (Schwingshackl et al., 2013) and the naturally occurring compound called Humulene(Satsu et al., 2004), can exclusively regulate the secretion of IL6 or IL8, respectively. The differential molecular regulation of IL6 and IL8 might be responsible for severe/moderate versus mild COVID-19 progression. Critical molecular regulators of IL6 and IL8, if identified, can provide a better hold on this disease. Several open questions need to be resolved for this. Are the levels of Trek-1 differential between severe, moderate, or mild patients? IL6 expression is regulated by TLRs and other PRR family mainly via NFkB signaling(Liu et al., 2017), nuclear factor of activated T cell (NFAT)(Nilsson et al., 2007), or Activator protein-1 (AP-1)(Ji et al., 2019). Are the regulators of these signaling arms differentially expressed in progressive patients? or are the suppressor of cytokine signaling(Verboogen et al., 2019) differential? Is IL6 upregulation more consequential than IL8-upregulation? If yes, Why? Also, we notice that all patients with severe disease manifestation and even some moderate patients show >50% lung damage, despite this lung specific biomarkers (SP-D or CC-16) are not elevated in the serum of IL6-low category of patients (Figure 6F compare dark-pink line-graphs with light-pink and green line-graphs). Does this represent specific type of lung damage via IL6 and/or other associated cytokines or is it due to some pre-existing lung condition? We have also noticed that few patients do not show any lung damage, using X-ray, but have moderate disease with high IL8-levels (>200pg/ml) (Supplementary Table1). What leads to oxygen deprivation in such patients? These preliminary observations need closer investigation(s). Initially, it was thought that the severe manifestation of this disease is due to age-mediated changes in the molecular profiles or due to co-morbidities. However, in the second wave of COVID-19 infection, these correlations did not hold, suggesting a constantly evolving picture. A lot still needs to be done at the molecular level to understand the molecular changes responsible for severe disease progression in patients, independent of their biological age.

Constantly mutating genome of SARS-CoV2, the increasing infectivity and their severity makes it challenging to act and think ahead of this virus. It is increasingly evident that many COVID-19 infected patients enter the progressive phase of the diseases due to virus-induced host immune reactions. Our study suggests that IL6 and IL8 levels must be considered for molecular stratification of COVID-19 patients, irrespective of their clinical stratification to pre-empt “cytokine-storm” dependent disease progression. Currently, most countries manage severe conditions only upon the onset of the symptoms, making it challenging to treat severe patients effectively to improve outcomes. IL6 and IL8 levels, rather than CRP levels, should be monitored to identify severe or moderate patients from mild for necessary and timely intervention with available immuno-modulators or precision drugs. Importantly, patients with IL6-low (<300pg/ml) and IL8-low (<200pg/ml) might be progressive due to direct virus-induced damage and should not be subjected to further immune suppression via immuno-modulatory drugs or precision medicines. It might be useful to evaluate if this subset of patients could rather benefit from antivirals drugs. Currently, based on the initial findings(Group et al., 2021), all patients with hypoxia (moderate or severe disease) are administered Dexamethasone (Supplementary Table 1). Our findings suggests that this current clinical practice needs a closer and critical evaluation. As we learn to live with this virus and its ever changing infectivity and severity pattern, resolving our clinical approach further will be useful in improving outcomes. However, our proposed IL6-high and IL8-high based patient stratification needs a larger study for validation and determination of a more precise cut-off for the cytokines based stratification. We envisage that our proposed stratification based on cytokine signatures may not be limited to SARS-CoV2 infections only. It might cover a larger canvas of viral or bacterial infectious agents known for causing cytokine-storm mediated organ damage and deaths. Nonetheless, this proposition needs further experimental validation.

## Methods and materials

### Samples

Venous blood was collected from PCR positive COVID-19 patients in EDTA-vaccutainers and clotting tube (1ml each). All blood samples were immediately stained for cell surface markers for FACS based analysis. Sample aliquoting as well as staining was done in BSL2 cabinet with PPEs. Serum samples were aliquoted and stored at -80°C freezer for subsequent ELISA based analysis. COVID-19 safety guidelines and protocols were followed for all wet lab based work using patient samples.

### Cell Staining for Flow Cytometric analysis

The antibody capture beads Unstained beads (Negative beads and Positive beads), single stained(negative beads + positive beads) were used for color compensation as per manufacturer’s protocol (552843 BD^™^ CompBead, Anti-Mouse). Antibody mix was prepared using 5 different antibodies according to their respective recommended concentrations. Precisely 50ul blood was added to it in 5 ml FACS™ tubes with antibodies. It The tubes was were incubated for 15 min in dark at room temperature. To lyse RBC, 450ul of 1X BD FACS™ lysing solution was added and again incubated for 15 min at room temperature. Antibody mix includes CD45 (563204, Mouse, BD Biosciences), CD38 (555462, Mouse, BD Biosciences), CD24 (Mouse, 555427, BD Biosciences), CD66B (Mouse, 561650, BD Biosciences), CD19 (Mouse, 560728, BD Biosciences). No spin Lyse no wash protocol was followed to avoid aerosol generation during sample processing. Samples were processed in BSL2 cabinet with appropriate PPEs.

### Enumeration of immunocytes using BD TruCount™ tubes by Flow Cytometry

Cell Staining: 10ul of individual antibodies were added in to BD TruCount ™ tubes (Catalog No. 340334) containing the beads according to their respective recommendations to product insert. Precisely, 50ul blood was added to it. It was incubated with recommended concentration of different antibodies for 15 min in dark at room temperature. To lyse RBC, 450ul of 1X BD FACS™ lysing solution was added and again incubated for 15 min at room temperature. Antibodies mix includes CD45, CD3, CD4, CD8 (Mouse, 340499, BD Biosciences). Samples were processed in BSL2 cabinet with appropriate PPEs. The scatter and fluorescence data of unstained and stained cells were acquired on a BD FACS Fortessa X-20™ Low cytometer using FACS™ diva software version V.8.0.3 (San Jose, USA).

The gating strategy applied to quantify the percentage of CD45, CD38, CD24, CD66B and CD19 cells is given in Supplement Figure 3. The absolute counts were obtained according to the BD TrueCount™ Tube product insert and gating strategy applied to enumerate the absolute number of each cell type is given in Supplement Figure 4. For Eg: To enumerate absolute count of the cell population (A), by dividing the number of positive cell events (X) by the number of bead events (Y), and then multiplying by the BD TruCount™ bead concentration (N/V, where N = number of beads per test* and V = test volume). A = X/Y × N/V.

### ELISA

Serum samples were diluted 1:5 times for ELISA analysis. Standards were prepared according to the manufacturers protocol. ELISA kits were used to prepare the samples according to each antibody. Cytokines analyzed include IL2 (0017212, BD Biosciences), IL6, (550799, BD Biosciences), IL8 (550999, BD Biosciences), IL4(550614, BD Biosciences), TNF-a (550610, BD Biosciences), and IL10 (550613, BD Biosciences) and Lung markers include Uteroglobin (DUGB00, R&D Systems), SP-D (DSFPD0, R&D Systems) and sRAGE (DRG00, R&D Systems). Samples were processed in BSL2 cabinet with appropriate PPEs.

## Data Availability

All data produced in the present work are contained in the manuscript

https://www.notapplicable.com

## Acknowledgements

NV and Anusha would like to acknowledge Dr. Mary Dias, Dr. Sivananda from IDU division for access to lab facilities. NCBS-Central Imaging and Flow Facility for FACS analysis. Mr. Bharat, NCBS, for help. Ms. Priya and Ms. Shakeela, Department of Medicine, for help with clinical data. Mr. Philip, SJMCH, for sample transportation. Anusha would like to thank Nitish Tayal and Nazia Chaudhary for their constant help. NV would like to acknowledge Prof. Apruva Sarin for her inputs on the study.

## Regulatory approvals

The study was approved by the Institutional Ethics Commitee at St. John’s Medical college and hospital (IEC ref. No. 144/2020, dated 22^nd^ May 2020). This project was reviewed by RCGM for Biosafety approval, DBT, GoI (on 27^th^ July 2020) and approved for the required wet-lab work with necessary safety compliances (Ref no. BT/17/006/96-PID, dated 2^nd^ July 2020).

## Informed consent

Venous blood and serum samples of Patients were collected by clinical team on the day of admission and at 48hrs, after taking written and informed consent.

## Authors contribution

Anusha = performed all the wetlab assays, provided wetlab data and manuscript draft, reviewed manuscript.

SAS = responsible for clinical data accrual, clinical data accuracy, patient classification, patient consent and reviewed manuscript.

RN = provided clinical samples, patients clinical data, patient consent.

BK = provided clinical samples from ICU.

GD = nucleated the team, designed the study and reviewed manuscript.

RK and HK = FACS data acquisition and analysis, reviewed manuscript.

JI = nucleated the clinical team, IERB approval, patient consent, overall clinical sample and team management, reviewed manuscript.

NV = designed the study, collected regulatory approvals, supervised the wet-lab assays, complied the clinical and wet-lab data, made figures, wrote manuscript.

