## Supplementary Figures for "Molecular markers for early stratification of disease severity and progression in COVID-19"

**Abstract**

COVID-19 infections have imposed immense pressure on the healthcare system of most countries. While the initial studies have identified better therapeutic and diagnostic approaches, the disease severity is still assessed by close monitoring of symptoms by healthcare professionals due to the lack of biomarkers for disease stratification. In this study, we have probed the immune and molecular profiles of COVID-19 patients at 48-hour intervals after hospitalization to identify early markers, if any, of disease progression and severity. Our study reveals that the molecular profiles of patients likely to enter the host-immune response mediated moderate or severe disease progression are distinct even in the early phase of infection when severe symptoms are not yet apparent. Our data from 37 patients suggest that at hospitalization, IL6 (>300pg/ml) and IL8 levels (>200pg/ml) identify cytokine-dependent disease progression. Monitoring their levels will facilitate timely intervention using available immunomodulators or precision medicines in those likely to progress due to cytokine storm and help improve outcomes. Additionally, it will also help identify cytokine-independent progressive patients, not likely to benefit from immuno-modulators or precision drugs.

*** Correspondence:**
Dr. Jyothi Idiculla:
Department of Medicine
St. John's Medical College and Hospital, SJNAHS.
Bangalore - 560 034. Karnataka, India.
Telephone No: +91 80 22065834


Dr. Neha Vyas:
Molecular Medicine Department
St. John's Research Institute, SJNAHS.
Bangalore - 560 034. Karnataka, India.
Telephone No: +91 80 49467125


Supplementary Figure 1: Clinical classification of COVID-19 patients

| Clinical Severity | Clinical presentation | Clinical parameters | Remarks |
|---|---|---|---|
| **Mild**[2] | Patients with uncomplicated upper respiratory tract infection, may have mild symptoms such as fever, cough, sore throat, nasal congestion, malaise, headache | Without shortness of breath or Hypoxia (normal saturation). | (i) Managed at Covid Care Centre OR at home (as per home isolation guidelines)[2] |
| **Moderate** | Pneumonia with no signs of severe disease | **Adults** with presence of clinical features of dyspnea and or hypoxia, fever, cough, including $SpO_2$ 90 to ≤93% on room air, Respiratory Rate more or equal to 24 per minute. | Managed in Dedicated Covid Health Centre (DCHC) |
| [illegible] | [illegible] | [illegible] | [illegible] |
| **Severe** | Severe Pneumonia | **Adults** with clinical signs of Pneumonia plus one of the following; respiratory rate >30 breaths/min, severe respiratory distress, $SpO_2$ <90% on room air. | Managed in Dedicated Covid Hospital (DCH) |
| | Acute Respiratory Distress Syndrome | **Onset**: new or worsening respiratory symptoms within one week of known clinical insult. **Chest imaging** (Chest X ray and portable bed side lung ultrasound): bilateral opacities, not fully explained by effusions, lobar or lung collapse, or nodules. **Origin of Pulmonary infiltrates**: respiratory failure not fully explained by cardiac failure or fluid overload. Need objective assessment (e.g. echocardiography) to exclude hydrostatic cause of infiltrates/ oedema if no risk factor present. **Oxygenation impairment in adults**: Mild ARDS: 200 mmHg < PaO2/FiO2 ≤ 300 mmHg (with PEEP or CPAP ≥5 cm H2O) Moderate ARDS: 100 mmHg < PaO2/FiO2 ≤200 mmHg with PEEP ≥5 cm H2O) Severe ARDS: $PaO_2/FiO_2$ ≤ 100 mmHg with PEEP ≥5 cm $H_2O$) | |

Supplementary Figure 2: IL4 levels are not different between COVID-19 patients with varying severity

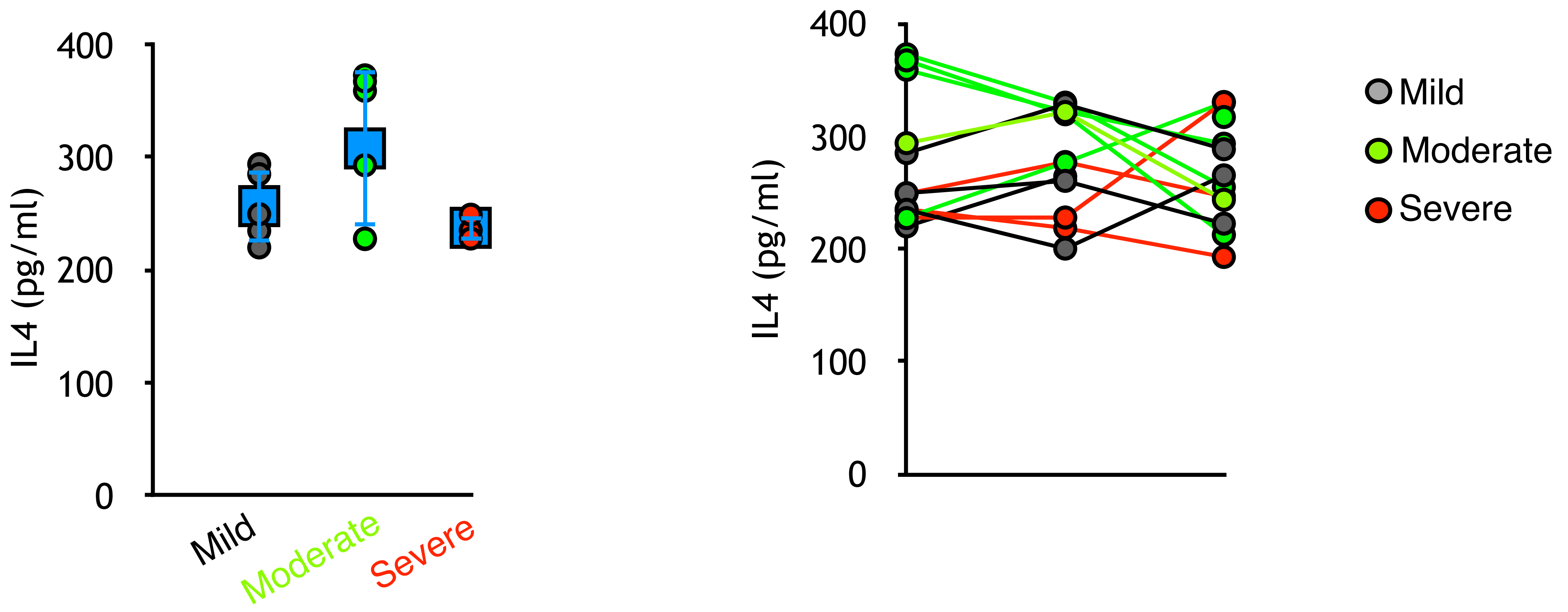

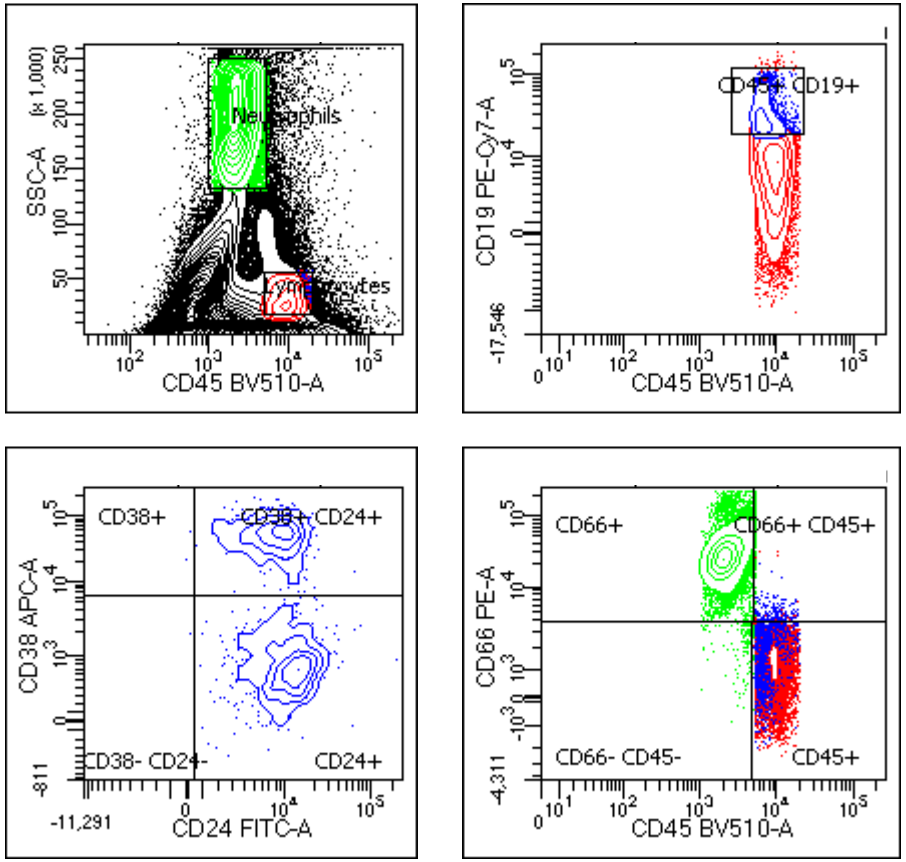


| Population | #Events | %Parent | %Total |
|---|---|---|---|
| All Events | 360,994 | #### | 100.0 |
| Lymphocytes | 6,252 | 1.7 | 1.7 |
| CD45+ CD19+ | 1,210 | 19.4 | 0.3 |
| CD38+ | 3 | 0.2 | 0.0 |
| CD38+ CD24+ | 460 | 38.0 | 0.1 |
| CD38- CD24- | 5 | 0.4 | 0.0 |
| CD24+ | 742 | 61.3 | 0.2 |
| Neutrophils | 38,182 | 10.6 | 10.6 |
| CD66+ | 37,900 | 99.3 | 10.5 |
| CD66+ CD45+ | 127 | 0.3 | 0.0 |
| CD66- CD45- | 132 | 0.3 | 0.0 |
| CD45+ | 23 | 0.1 | 0.0 |

Supplement Figure 3

The CD45 positive Lymphocytes and Neutrophils were selected on
CD45 and Side Scatter plot. The CD45 and CD19 positive cells gated on CD38 and CD24 plot. The Neutrophils were gated on CD66 and CD45 plot.

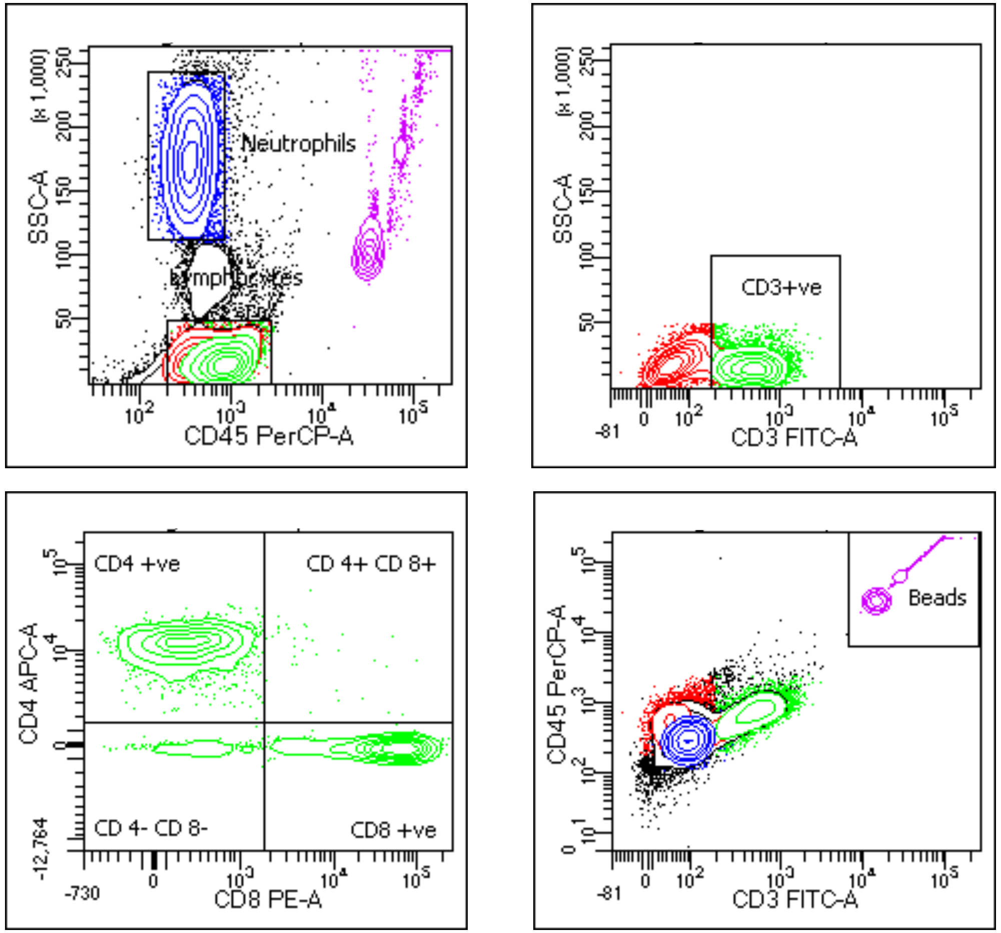


Supplement Figure 4

The CD45 positive Lymphocytes and Neutrophils were selected on
CD45 and Side Scatter plot. The CD45 and CD3 cells were gated on CD4 and CD8 plot. The TruCount bead population on CD3 and CD45 plot was considered for calculating absolute count of various populations.

| Population | #Events | %Parent | %Total |
|---|---|---|---|
| All Events | 20,000 | #### | 100.0 |
| Lymphocytes | 7,301 | 36.5 | 36.5 |
| CD3+ve | 3,575 | 49.0 | 17.9 |
| CD4 +ve | 1,946 | 54.4 | 9.7 |
| CD 4+ CD 8+ | 34 | 1.0 | 0.2 |
| CD 4- CD 8- | 221 | 6.2 | 1.1 |
| CD8 +ve | 1,374 | 38.4 | 6.9 |
| Neutrophils | 7,691 | 38.5 | 38.5 |
| Beads | 2,069 | 10.3 | 10.3 |
