## Supplementary Table 1 for "Molecular markers for early stratification of disease severity and progression in COVID-19"

### Abstract

COVID-19 infections have imposed immense pressure on the healthcare system of most countries. While the initial studies have identified better therapeutic and diagnostic approaches, the disease severity is still assessed by close monitoring of symptoms by healthcare professionals due to the lack of biomarkers for disease stratification. In this study, we have probed the immune and molecular profiles of COVID-19 patients at 48-hour intervals after hospitalization to identify early markers, if any, of disease progression and severity. Our study reveals that the molecular profiles of patients likely to enter the host-immune response mediated moderate or severe disease progression are distinct even in the early phase of infection when severe symptoms are not yet apparent. Our data from 37 patients suggest that at hospitalization, IL6 (>300pg/ml) and IL8 levels (>200pg/ml) identify cytokine-dependent disease progression. Monitoring their levels will facilitate timely intervention using available immunomodulators or precision medicines in those likely to progress due to cytokine storm and help improve outcomes. Additionally, it will also help identify cytokine-independent progressive patients, not likely to benefit from immuno-modulators or precision drugs.

*** Correspondence:**
Dr. Jyothi Idiculla:
Department of Medicine
St. John's Medical College and Hospital, SJNAHS.
Bangalore - 560 034. Karnataka, India.
Telephone No: +91 80 22065834


Dr. Neha Vyas:
Molecular Medicine Department
St. John's Research Institute, SJNAHS.
Bangalore - 560 034. Karnataka, India.
Telephone No: +91 80 49467125


| | Supplementary Table 1: COVID-19 Patients clinical details | | | | | | | | | | |
|---|---|---|---|---|---|---|---|---|---|---|---|
| | | | Treatment | | | | | | | | |
| | Research lab Serial no. | Age | Dexamethasone | Remedesivir | Tocilizumab | Chest X-ray (CXR) | Symptom to Hospitalization (days) | Hospital stay (days) | COVID category | O2 levels | Remark |
| Mild COVID | Patient 01 | 21-25 | Yes | No | No | b/l infiltrates <50% | 10 | 3 | UNEVENTFUL, MILD COVID | >94% | |
| | Patient 02 | 31-35 | No | No | No | Normal | 3 | 3 | MILD COVID | >94% | |
| | Patient 03 | 21-25 | No | No | No | NORMAL | 10 | 3 | MILD COVID | >94% | |
| | Patient 04 | 46-50 | No | No | No | NORMAL | 3 | 7 | MILD COVID | >94% | |
| | Patient 05 | ?? | No | No | No | NORMAL | 3 | 7 | UNEVENTFUL, MILD COVID | >94% | |
| | Patient 06 | 46-50 | No | No | No | NORMAL | 7 | 10 | MILD COVID | >94% | |
| | Patient-19 | 51-55 | No | No | No | NORMAL | 5 | 15 | MILD COVID | >94% | |
| | Patient 21 | 66-70 | No | No | No | 60% infiltrates | 4 | 12 | MILD COVID, UNEVENTFUL | >94% | |
| | Patient 32 | 66-70 | No | No | No | NORMAL | 20 | 6 | MILD COVID | >94% | |
| | Patient 09 | 71-75 | No | No | No | B/L MID AND LOWER ZONE OPACITIES WHICH WORSENED TRANSIENTLY FROM 30% TO 50% INVOLVEMENT AND IMPROVED AT DISCHARGE | 10 | 12 | MILD COVID | | |
| | Patient 41 | 26-30 | - | - | - | CXR : B/L infilterates 30% involvement | 1 | 4 | MILD COVID | >94% | **Not included** |
| | Patient 43 | 36-40 | - | - | - | Left diffuse infilterates 30-40% involvement | 15 | 5 | MILD COVID?? | >94% | **Not included** |
| Moderate COVID | PAtient 07 | 51-55 | Yes | No | No | MILD LOWER ZONE INFILTRATES ~50% INVOLVEMENT | 3 | 4 | MODERATE COVID | 90-93% | |
| | Patient 11 | 46-50 | Yes | Yes | No | Cxr: 60% lung involvement upper mid and loer zone opacities + | 4 | 6 | MODERATE COVID | 90-93% | **Not included** |
| | Patient 12 | 81-85 | Yes | No | No | NORMAL | 5 | 8 | MODERATE COVID | 90-93% | |
| | Patient 13 | 71-75 | Yes | No | No | CXR: LEFT LOWER ZONE INFILTRATES WITH CP ANGLE BLUNTIING 15-30 % INVOLVEMENT. | 3 | 24 | MODERATE COVID | 90-93% | |
| | PAtient 17 | 56-60 | Yes | No | No | CXR : Supine AP, poor expiratory film, apparent cardiomegaly, Bilateral infilterates with >50% involvement | 10 | 17 | IMPROVED, MODERATE COVID RECOVERED AND DISCHARGED, STAY UNEVENTFUL | 90-93% | |
| | Patient 18 | 41-45 | Yes | No | No | 40-50% infiltrates | 4 | 6 | MODERATE COVID, IMRPOVED WITH STEROIDS AND OXYGENATION | 90-93% | |
| | Patient 23 | 51-55 | Yes | No | No | MILD LOWER ZONE INFILTRATES, <50% | 3 | 12 | MODERATE COVID | 90-93% | |
| | Patient 25 | 56-60 | Yes | Yes | No | BILTERAL MINIMAL LOWER ZONE INFILTRATES 10-20% | 5 | 10 | MODERATE COVID | 90-93% | |
| | Patient 26 | 51-55 | Yes | No | No | CXR : B/L infilterates 40-50% involvement CT score - 12/25 Peripheral B/L ground glass opacities with consolidation. | 3 | 4 | MODERATE COVID | 90-93% | |
| | Patient 35 | 31-35 | Yes | No | No | CXR : B/L infilterates 30-40 % involvement | 20 | 5 | MODERATE COVID | 90-93% | |
| | Patient 38 | 46-50 | Yes | No | No | CXR : 30-40 % involvement | 8 | 5 | MODERATE COVID | **93%** | |
| | Patient 10 | 81-85 | Yes | No | No | NORMAL | 3 | 8 | MODERATE COVID | 90-93% | |
| | Patient 08 | 71-75 | Yes | No | No | CXR: B/L NON HOMOGENOUS OPACITIES 40-50% INVOLVEMENT | 10 | 10 | SEVERE COVID | **<90%** | |
| | Patient 14 | 21-25 | Yes | Yes | No | CXR: B/L UPPER MID AND LOWER ZONE OPACITIES 60% INVOLVEMENT | 4 | 14 | SEVERE COVID | **<90%** | |

| | | | | | | | | | | | |
|---|---|---|---|---|---|---|---|---|---|---|---|
| | Patient 15 | 56-60 | Yes | No | No | CT KUB (PLAIN) IMP:Bilateral bulky kidneys with perinephric and periureteric fat stranding, bilateral hydroureteronephrosis (R >L) with urothelial thickening - s/o Pyelonephritis. | 7 | 25 | SEVERE COVID | **<90% (83%)** | |
| | Patient 16 | 66-70 | Yes | Yes | No | CXR: 40% INVOLVEMENT MID AND LOWER ZONE INFILTRATES | 7 | 17 | SEVERE COVID | **<90%** | |
| | Patient 20 | 31-35 | Yes | No | No | EXTENSIVE GROUND GLASSING | 5 | Day6 shifted to ICU | SEVERE COVID | **<90%** | |
| | Patient 22 | 61-65 | Yes | Yes | No | EXTENSIVE SHADOWS 70-80% INVOLVEMENT | 7 | 25 | DIAGNOSIS: SEVERE COVID WITH POST COVID SEQUELAE | **<90%** | |
| | Patient 24 | 76-80 | Yes | Yes | No | HRCT: CORADS 6 ~70% LUNG INVOLVEMENT (16/25, CT) | 5 | 7th day, Death | SEVERE COVID | **<90%** | **Not included** |
| | Patient 27 | 41-45 | Yes | Yes | Yes | cxr : b/l Mid zone and lower zone infilterates 30-40 %, CT PA; on 23/6/21 revealing diffuse ground glass opacitiess with crazy paving in bilateral lung fields,CT involvementcore of 24/25. | 4 | 19 | SEVERE COVID | **<90%** | **Not included** |
| **Severe COVID** | Patient 28 | 66-70 | Yes | No | No | Chest Xray-B/L patchy infilterates 70% lung involvement | 1 | 6the day , Death | SEVERE COVID | **<90%** | |
| | Patient 30 | 36-40 | Yes | Yes | No | B/L costophrenic and left cardiophrenia angle blunting. Opacities on left lower and upper zone. Pleural effsuion minimal to moderate, left more than right. 60 - 70 % involvement | 4 | 11 | SEVERE COVID | **<90%** | |
| | Patient 33 | 36-40 | Yes | ? | ? | CXR : B/L diffuse infilterates with >90 % involvement, 26/26 HRCT | 2 | 28th Death | Severe COVID POST COVID ILLNESS. Death | **<90%** | |
| | Patient 34 | 41-45 | Yes | Yes | No | CXR : B/L infilterates right > left 60-70 % involvement | 4 | 8 | SEVERE COVID | **<90%** | |
| | Patient 36 | 61-65 | Yes | No | No | Right diffuse 25% infilterates 9/40 | 2 | 8 | SEVERE COVID | **<90%** | |
| | Patient 37 | 56-60 | Yes | Yes | Yes | CXR : B/L infilterates 90-95% involvement | 3 | 22nd Day death | SEVERE COVID | **<90%** | |
| | Patient 39 | 61-65 | Yes | Yes | No | CXR : B/L infilterates 80% | 6 | 33 | SEVERE COVID | **<90%** | |
| | Patient 40 | 76-80 | Yes | Yes | No | CXR : B/L infilterates with 60 % involvement c. CT - HRCT-21/6/21- Partial/chronic pulmonary thrombo embolism involving the left lower lobar subsegmenta,l branches., 24/25, HRCT, >90% infiltrates Features of typical COVID Pneumonia - CORADS 6 with CT involvement score of 24 /25 (severe). | 6 | 18 | SEVERE COVID | **<90%** | |
| | Patient 42 | 81-85 | Yes | No | No | CXR : diffuse infilterates 60-70%, 16/25 HRCT | 14 | 6 | SEVERE COVID | **<90%** | |
| | Patient 44 | 31-35 | Yes | Yes | No | CXR : B/L infilterates 30% | 10 | 54, Death | SEVERE COVID | **<90%** | |
